# Using Histopathology Images to Predict Chromosomal Instability in Breast Cancer: A Deep Learning Approach

**DOI:** 10.1101/2020.09.23.20200139

**Authors:** Zhuoran Xu, Akanksha Verma, Uska Naveed, Samuel Bakhoum, Pegah Khosravi, Olivier Elemento

## Abstract

Chromosomal instability (CIN) is a hallmark of human cancer that involves mis-segregation of chromosomes during mitosis, leading to aneuploidy and genomic copy number heterogeneity. CIN is a prognostic marker in a variety of cancers, yet, gold-standard experimental assessment of chromosome mis-segregation is difficult in the routine clinical setting. As a result, CIN status is not readily testable for cancer patients in such setting. On the other hand, the gold-standard for cancer diagnosis and grading, histopathological examinations, are ubiquitously available. In this study, we sought to explore whether CIN status can be predicted using hematoxylin and eosin (H&E) histology in breast cancer patients. Specifically, we examined whether CIN, defined using a genomic aneuploidy burden approach, can be predicted using a deep learning-based model. We applied transfer learning on convolutional neural network (CNN) models to extract histological features and trained a multilayer perceptron (MLP) after aggregating patch features obtained from whole slide images. When applied to a breast cancer cohort of 1,010 patients (Training set: n=858 patients, Test set: n=152 patients) from The Cancer Genome Atlas (TCGA) where 485 patients have high CIN status, our model accurately classified CIN status, achieving an area under the curve (AUC) of 0.822 with 81.2% sensitivity and 68.7% specificity in the test set. Patch-level predictions of CIN status suggested intra-tumor spatial heterogeneity within slides. Moreover, presence of patches with high predicted CIN score within an entire slide was more predictive of clinical outcome than the average CIN score of the slide, thus underscoring the clinical importance of spatial heterogeneity. Overall, we demonstrated the ability of deep learning methods to predict CIN status based on histopathology slide images. Our model is not breast cancer subtype specific and the method can be potentially extended to other cancer types.

## Introduction

Chromosomal instability (CIN) refers to ongoing chromosome segregation errors throughout consecutive cell divisions that can potentially result in extensive numerical and structural chromosomal aberrations [1] [2]. CIN, as one of the hallmarks of human cancer, has been recognized as a central driver of cancer evolution owning to its multipronged effects which facilitate processes such as metastasis, immune evasion and therapeutic resistance [3, 4]. For example, Smid et al. [5] found that elevated CIN and consequent high aneuploidy burden is associated with poor breast cancer prognosis, measured as time to distant metastasis. Carter et al. [6] revealed a correlation between a transcriptional signature of CIN with metastasis, tumor grading and clinical outcome in multiple human cancers. Given the widespread nature and far-reaching consequences of CIN in human cancer, strategies for targeting CIN as a therapeutic vulnerability are being actively researched [7] [8]. Despite its clear importance, CIN status is not readily testable for cancer patients in routine clinical settings because it requires complicated experimental assessment either involving live microscopy, sensitive detection of micronuclei (a consequence of CIN) via immunohistochemistry or comprehensive genomic analysis. On the other hand, gold-standard histopathological examinations that are used for cancer diagnosis and grading are ubiquitously available. Here we sought to investigate the feasibility of using histopathology whole slide images to predict CIN status.

Deep learning is a state-of-the-art methodology for analyzing and interpreting cancer histology images. In recent years, a large number of studies attempted to employ deep learning approach for a variety of tasks in computational pathology field by taking advantage of its ability to extract hierarchical features from images in a direct and automatic fashion. Previous research has shown that presence of driver mutations, mutational signatures and expression-defined tumor subtypes can be predicted from H&E slides [9, 10, 11]. For example, Kather et al. [12] trained a Convolutional Neural Network (CNN) model that can robustly predict genome microsatellite instability (MSI) in gastrointestinal cancer from H&E histology, obtaining patient level area under the curve (AUC) of 0.84. Coudray et al. [9] trained deep learning network that successfully predicted six out of ten most commonly mutated genes from lung adenocarcinoma (LUAD) pathology images, with AUCs from 0.733 to 0.856. Kather et al. [13] then demonstrated the ability of deep learning to predict point mutations, molecular tumor subtypes and immune-related gene expression signatures directly from H&E images in multiple cancer types. Fu et al. [14] used transfer learning and correlated histopathological pattern features with genomic, transcriptomic and survival data in 28 cancer types.

In this study, we proposed using pathology images to predict patients’ CIN status in breast cancer. We created a framework that uses transfer learning and feature aggregation to accurately discriminate high CIN and low CIN histopathology slides without human intervention. Our results indicate that (1) CIN can be predicted accurately from histopathology slides (2) unexpectedly there appears to be substantial spatial heterogeneity CIN status in many patients. These results pave the way for using CIN as a biomarker of prognosis and response to anti-CIN therapies fully integrated into existing clinical pathology workflows.

## Results

### A weakly supervised deep learning model for patient genomic CIN classification in breast cancer

We obtained H&E slides and genomic profiles from breast cancer patients in TCGA. Here we quantify CIN using the fraction genome altered (FGA, see Methods). FGA quantifies the burden of aneuploidies detectable in bulk genomic profiles. While not a perfect measure of CIN, we and others have observed a strong correlation between FGA and CIN measured using microscopy and/or micronuclei staining [15]. We refer to FGA as genomic CIN score, to contrast it with pathology predicted CIN scores introduced in this study. Genomic CIN score higher than 0.3 was labelled as high CIN; genomic CIN score lower than 0.3 was labelled as low CIN (**Figure S1**). H&E slides were processed as described in Methods. Based on our CIN classification, the breast cancer (BRCA) cohort had 485 high CIN patients with 515 WSI and 23,427 patches, and 525 low CIN patients with 550 WSI and 23,568 patches (**Table S1**). This study presents a deep learning model that can automatically predict patients’ genomic CIN status on molecular level from H&E stained histopathology slides (**Figure 1a-e**, see Methods). Our model uses Convolutional Neural Network (CNN) models pre-trained on ImageNet as patch-level feature extractor (**Figure 1b**) and then aggregated patch features into patient features (**Figure 1c**). This approach effectively addresses intra-tumor heterogeneity using “weak” patient-level labels, but also offers opportunity to explore spatial CIN heterogeneity in individual patients (**Figure 1d-e**). The 1,070 Whole Slide Images (WSI) of 1,010 patients from TCGA-BRCA were randomly split into training, validation and test set. We then evaluated model performance in the hold-out test set, which included 152 patients.

**Figure 1.**
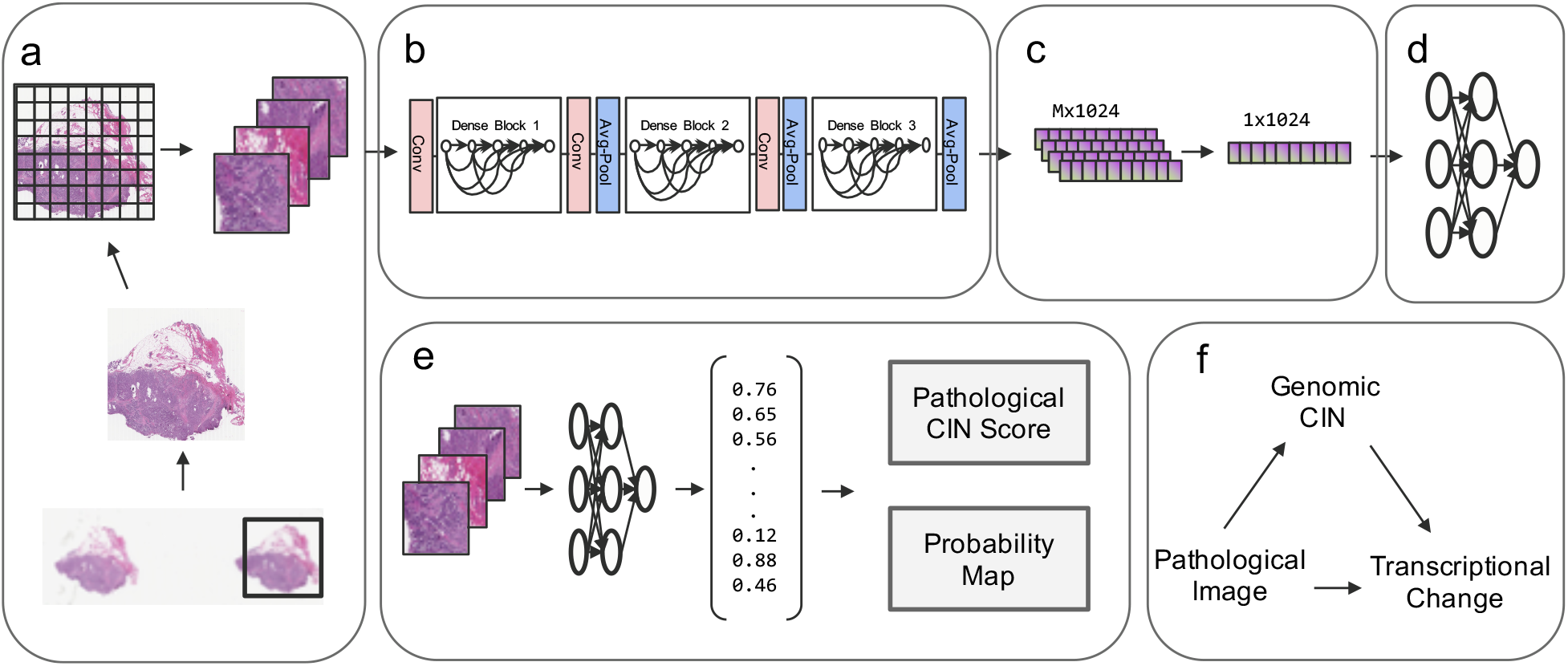
Overview of the pipeline. **(a)** During mage preprocessing, an overall ROI was identified in WSI as window with highest tissue percentage using a sliding window approach. Then the ROI was tiled into non-overlapping patches before quality control. Only qualified patches were kept as described in Methods. **(b)** Patch-level feature extraction was performed using a pre-trained CNN architecture. **(c)** Max-pooling layer was used for aggregating patch level features to patient level features. **(d)** MLP was trained in supervised approach based on patient-level genomic CIN status. **(e)** Patch level features were fed into trained MLP in d. Pathology predicted CIN scores were calculated and probability maps based on patch predictions were generated. **(f)** Pathological Images were used to predict genomic CIN. Since genomic CIN can potentially alter gene expression and pathways on transcriptional level, differentially expressed gene analysis and pathway analysis were performed. Pathology predicted CIN and Genomic CIN were compared to CIN transcriptional signatures.

### Deep learning model predicts CIN with high accuracy and sensitivity

Several commonly used CNN architectures were tested in the transfer learning step used to extract the most relevant features that can predict genomic CIN. The best feature extraction method was selected based on ability of trained MLP to predict CIN groups. Results shown in **Figure 2** indicate that Densenet-121 achieved the best performance with AUC of 0.822 and accuracy of 74.3%. Densenet networks with different depth achieved similar performance with AUC of 0.806 and 0.807 for Densenet-169 and Densenet-201 respectively. The Densenet-121 model got a good sensitivity of 81.16%. Xception model achieved AUC of 0.752 and Resnet-50 with 0.650. Altogether these results indicate that a deep learning model can accurately classify CIN status, achieving an area under the curve (AUC) of 0.822 with 81.2% sensitivity and 68.7% specificity in an independent test set not used for training or parameter exploration.

**Figure 2.**
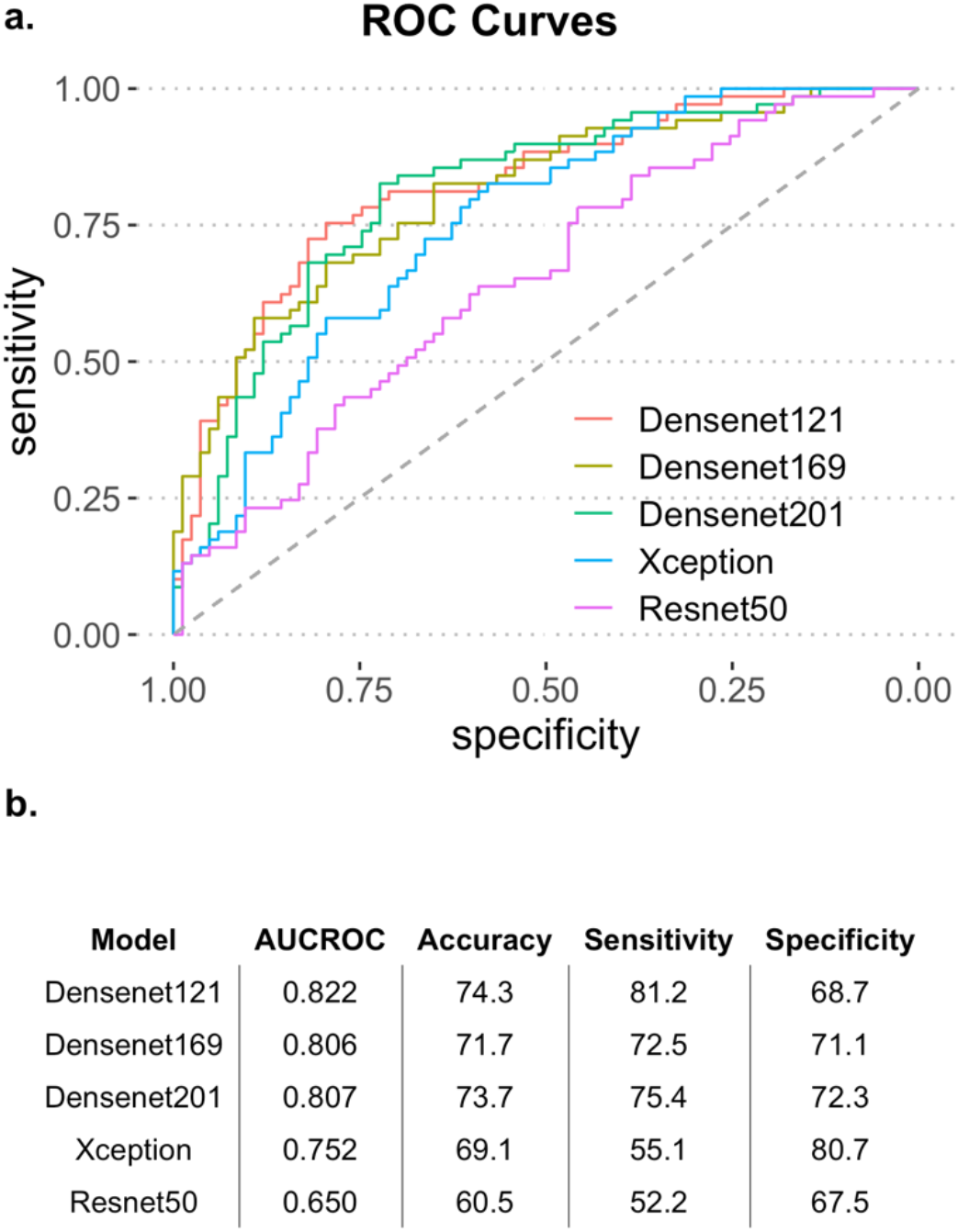
Model performance final evaluation. Model performance was evaluated in test set. **(a)** ROC curves for different CNN architectures. **(b)** Table for model performance with AUC, accuracy, sensitivity and specificity.

### High CIN patients exhibits more atypical mitosis events

To independently validate that pathology-predicted CIN status (and genomic CIN) correlates with CIN related aberrant mitotic events, we inspected 10 tumor slides at 40x magnification level, looking manually for aberrant mitotic events. Half of the 10 slides were predicted as high CIN and half as low CIN. All 10 slides were also concordantly labelled as high CIN or low CIN by genomic CIN (in other words, they were true positive and true negative predictions). As shown in **Figure 3a-h**, both normal and abnormal mitosis events including anaphase bridge, spindles with misalignment chromosomes, multipolar and monopolar chromosome arrangements were observed. By fitting Generalized estimating equation (GEE) Poisson regression model, we found that atypical mitosis event counts per field of view (patches with fixed size of 1,024*1,024 pixels on 40x) are significantly higher in predicted high CIN patients comparing with predicted low CIN patients (p-value<0.0001, **Figure 3i**).

**Figure 3.**
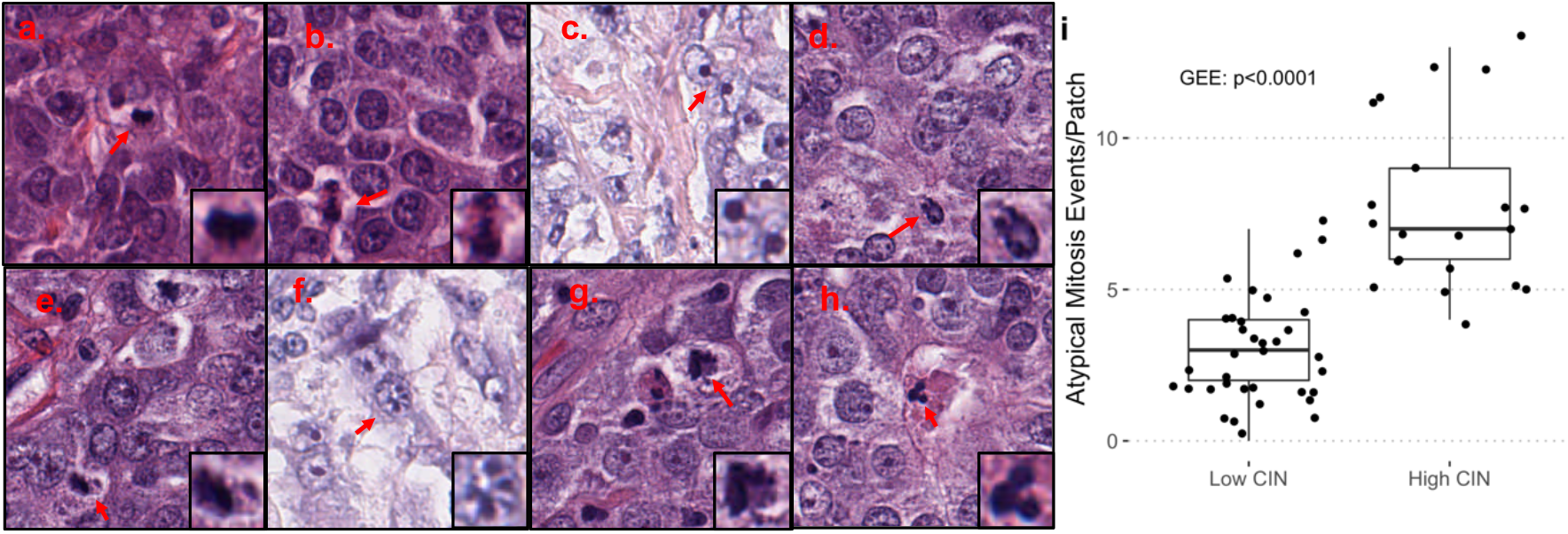
Patches containing normal and abnormal mitosis events at 40x magnification. **(a)-(b)** Normal mitosis; **(c)-(h)** Abnormal mitosis. **(c)** Anaphase bridge. **(d)** Monopolar mitosis. **(e)-(g)** Mitotic figure with unaligned chromosomes. **(h)** Multipolar mitosis. **(i)** Boxplot of atypical mitosis events per field of view between 5 predicted high CIN and 5 predicted low CIN patients.

### Patch predictions demonstrates intra-tumor heterogeneity

As discussed in the previous section, we had hypothesized that not all patches within the same slide may have the same level of CIN; in other words, we hypothesized that there may be spatial heterogeneity in CIN within the same tissue section. To test this hypothesis, we visualized patch predictions within one WSI. Patch predictions were generated by feeding individual patch features into trained MLP independently. As shown in **Figure 4**, both high CIN and low CIN patches can be found within one WSI regardless of the slide’s CIN status. **Figure 4a** is an example of low CIN slide with predicted high CIN probability of 0.27. Out of all 45 patches, 11 (= 24.44%) were predicted as high CIN patches. Because high CIN patches still exist in low CIN slides due to intra-tumor heterogeneity, the prevalence scale and patch probability also have influence of the whole slide CIN status. Similar results were shown in **Figure 4b**, where low CIN patches were also found in high CIN slide. We thus define predicted CIN-high fraction score as the percentage of predicted high CIN patches based on each pathology image. Altogether we found that only 94 (9.31%) out of 1010 patients in the whole cohort exhibited low level of spatial heterogeneity with predicted CIN-high fraction score smaller than 10%. The median predicted CIN-high fraction score of this cohort is 57% with 25^th^ and 75^th^ percentile of 32% and 83% respectively (**Figure S2**).

**Figure 4.**
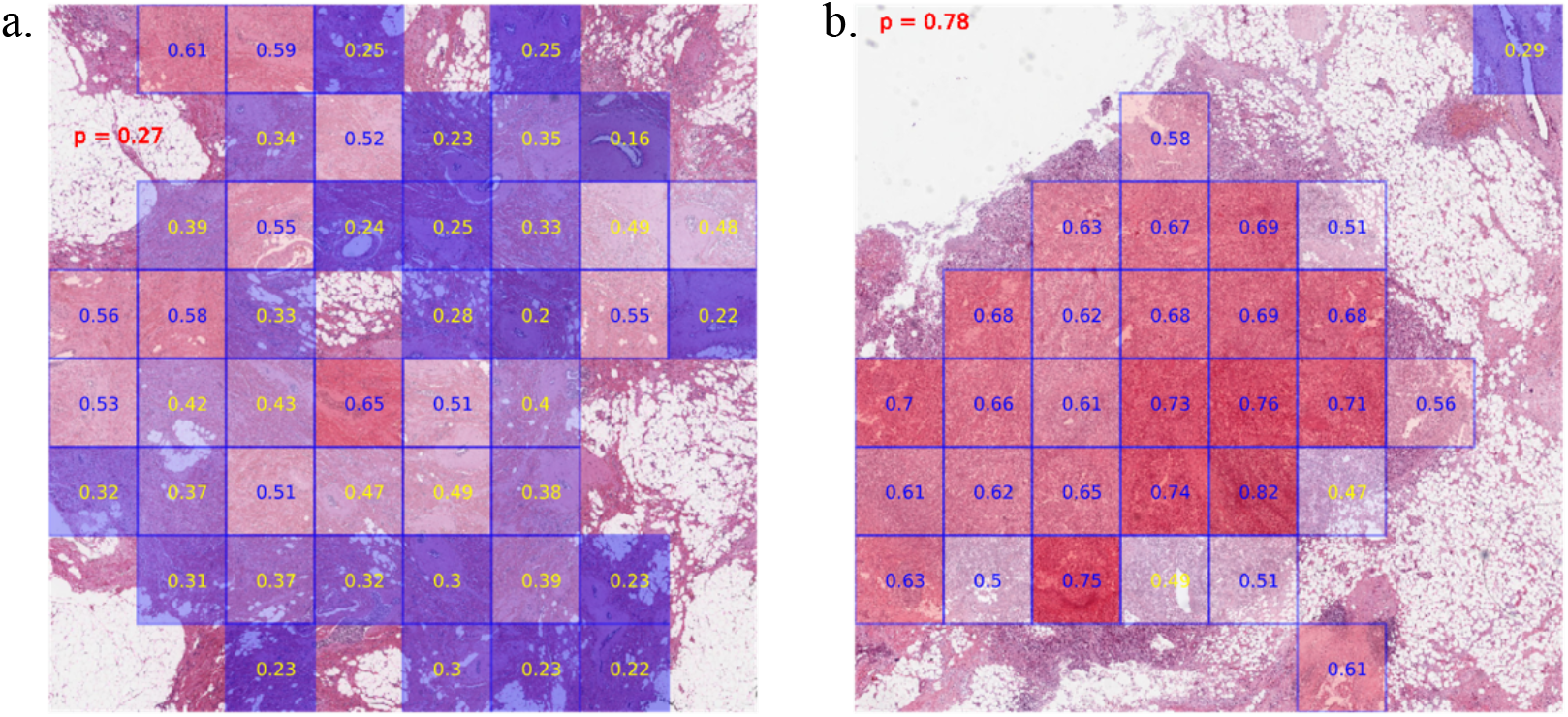
Intra-tumor heterogeneity by patch predictions. Red shows predicted probability of high CIN slide based on our deep learning model. Numbers within each grid imply patch level predictions. Blue indicates high CIN patch, yellow indicate low CIN patch. **(a)** an example of low CIN slide in test set. **(b)** an example of high CIN slide in test set

### Fraction of predicted CIN high patches is correlated with Transcriptional CIN score

CIN, as a hallmark of cancer has been linked to activation of key downstream biological pathways such as cGAS-STING and non-canonical NF-kb. [3] To bridge the gap between molecular genome alternations with pathological features, we conducted correlation analysis between a CIN driven transcriptional gene signature with both genomic CIN score and predicted CIN-high fraction score. CIN23 is a gene signature derived from the human metastatic cell line models (MDA-MB-231) engineered to over-express MCAK (to suppress CIN) or a dominant negative version of MCAK (to increase CIN) [4]. Employing CIN23, we derived a gene signature score for each patient as transcriptional CIN score using ssGSEA. Genomic CIN score was positively correlated with transcriptional CIN score with correlation coefficient of 0.14 (**Table S4**, p-value<0.0001). We then reasoned that the average predicted CIN spatial heterogeneity measured by the percentage of predicted high CIN patches of each slide may correlate with the transcriptional CIN score, which is also an average representation of CIN across all spatial areas. Indeed, we observed a weak but significant positive correlation with transcriptional CIN (**Table S4**, rho: 0.1, p-value=0.0026).

### Model performance is not breast cancer subtype specific

In the past, different patterns of CIN were observed to be associated with distinct subtypes of breast cancer. [16] In this cohort, we found significant positive association between genomic CIN status with the prevalence of ER-(p-value<0.0001), PR-(p-value<0.0001) and Triple Negative (p-value<0.0001) subtype status but not HER2 (p-value=0.1) status (**Table S2**). We therefore sought to verify that our algorithm is not simply predicting tumor subtypes (since we have that predicting breast cancer subtypes is feasible from H&E slides [17]). We combined subtype information (ER, PR, HER2 status) along with clinical features including age, race, menopause status and number of positive lymph-nodes with image input and retrained an MLP model. Adding these features did not improve the model performance (p-value=0.82) (**Fig S3**). Finally, no evidence suggested our model to be subtype specific with AUCs statistically the same across different subtypes (p-values: ER: 0.33, PR: 0.94, HER2: 0.41, Triple Negative: 0.86) in this TCGA cohort (**Table S3**). We concluded that our model is predictive of CIN independently of tumor subtypes.

### CIN is associated with poor prognosis in breast cancer

The association between CIN and cancer prognosis is complex and paradoxical. Some studies showed association of CIN with poorer cancer prognosis [18, 6], while other studies reached opposite conclusions, suggesting that excessive level of CIN would suppress tumor progression and lead to better clinical outcomes [19, 20]. To further investigate this point, we conducted a survival analysis in the TCGA cohort aiming to explore the prognostic values of different CIN scores. In these analyses, we used maximally selected rank statistics to determine optimal prognostic CIN score cutoffs and log-rank tests to evaluate the differences between survival curves. We found that high genomic CIN is associated with poorer 5 years’ prognosis compared to low genomic CIN, where prognosis is measured as time to any events including new tumors or mortality (**Figure 5a**. p-value=0.0023). Pathology-predicted CIN using our deep learning model was also predictive of outcomes (**Figure 5b**. p-value=0.0045). Finally, predicted CIN-High fraction score was also correlated with worse outcomes (**Figure 5c**. p-value=0.0086). We postulated that the presence of patches with high predicted CIN scores within each slide may be sufficient to impact clinical outcomes. We calculated different percentile CIN scores based on all patches of each slide (75^th^, 95^th^ and maximum). We found that all slide-level percentile CIN scores were prognostic (**Figure 5d-f**. p-values: CIN-75^th^: 0.0085, CIN-95^th^: 0.0018, CIN-max: 0.02) with CIN-95^th^ being the most robust prognostic score, even more predictive than genomic CIN. That CIN-95^th^ is more predictive than CIN-max can be explained by the lack of stability of maximal value and possibly by the need for more than one patches to have high CIN to impact outcomes.

**Figure 5.**
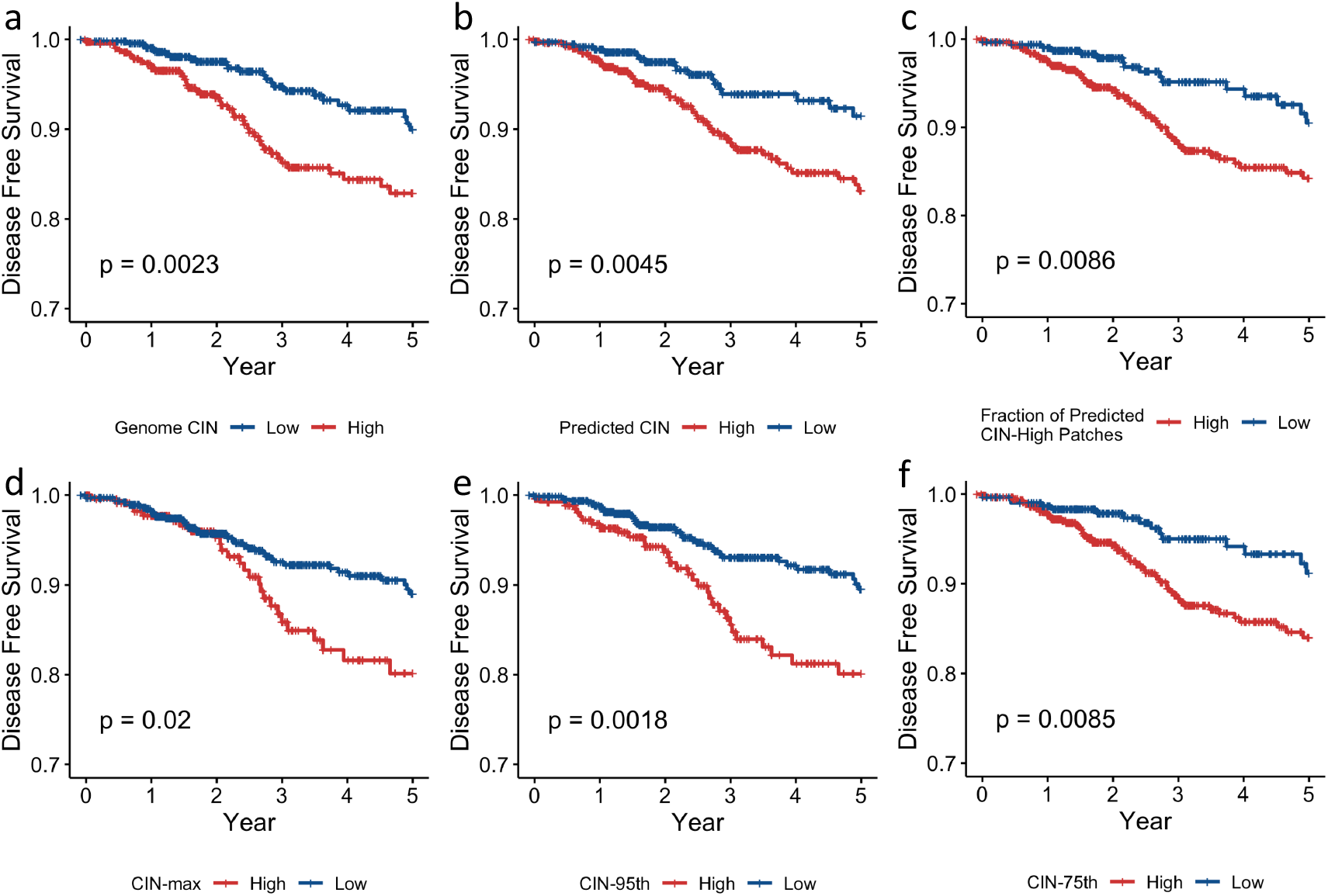
Kaplan-Meier curves of disease-free survival (DFS) probabilities grouped by different CIN biomarkers. P values were calculated by log rank test. **(a)** Genomic CIN (Stratification cutoff: 0.3). **(b)** Predicted CIN (Stratification cutoff: 0.44). **(c)** Fraction of predicted CIN high patches (Stratification cutoff: 0.42). **(d)** CIN-max, the maximum patch prediction within each slide (Stratification cutoff: 0.77). **(e)** CIN-95^th^, the 95^th^ percentile patch prediction within each slide (Stratification cutoff: 0.72). **(f)** CIN-75^th^, the 75th percentile patch prediction within each slide (Stratification cutoff: 0.55).

### CIN is associated with profound transcriptional changes in tumor samples

We reasoned that our ability to predict CIN based on histopathology slides may underlie a relatively profound difference in biological features between CIN low and CIN high tumors, which may influence cell morphology and tissue structure in H&E slides. We therefore conducted differentially expressed genes and gene set enrichment analysis between high CIN and low CIN samples. To minimize the confounding effect of cancer subtypes caused by the unbalanced subtype distributions across CIN groups, we adjusted tumor subtypes in the design matrix and tested differentially expressed genes for genomic CIN term. 307 differentially expressed genes were identified (logFC>1, adjusted p-value<0.05) between CIN low and CIN high tumors as shown in **Figure S8a**. Cell cycle and mitosis related gene signatures were up-regulated significantly in high CIN tumor samples (**Figure S8b**). This analysis thus confirms substantial biological differences between CIN high and CIN low tumors.

## Conclusion and Discussion

In this study we demonstrate for the first time the ability to predict chromosomal instability (CIN) based on H&E slides. Currently it is challenging to capture the ongoing rate of chromosome mis-segregation to identify CIN in routine clinical setting since mitotic alterations are rare in H&E slides; other assays such as microscopy or micronuclei staining have not been deployed in the clinical setting. Here we demonstrated the ability of using histopathology slide images to predict CIN status of each patient and achieved high accuracy (=74.3%) and sensitivity (=81.2%). Equally importantly, our model indicates the existence of intra-tumor spatial heterogeneity in CIN levels and revealed its association with poorer clinical outcomes. Further research, perhaps based on regional sequencing, is needed to further validate these findings. The substantial prognostic impact potentially exerted by spatial sub-regions (patches) with highest CIN scores is important since it indicates that such regions may drive response to treatment. Future treatment modalities may need to focus on eliminating CIN high tumor cells if they are to achieve maximal therapeutic impact. Either way, our results pave the way for integrating CIN as prognosis biomarker and therapeutic vulnerability into existing clinical pathology settings.

One of the main challenges of computational pathology is to manage the tradeoff between abundant morphological information and large size of whole slide image. Splitting WSI into hundreds of thousands of patches and training neural networks on patch level is a commonly used strategy [21]. As mentioned above, there are several studies that successfully demonstrated the ability of using H&E stained histology to predict genetic mutations using this patch-level learning approach [17, 12]. We also experimented using patients’ level labels to supervise patch learnings directly with the same approach but failed in predicting CIN levels. We reasoned there might exist substantial intra-tumor heterogeneity within individual slides, and that therefore using patient level CIN labels are not directly applicable to patches for training. To overcome this obstacle for CIN status learning, we applied a weakly supervised learning approach. More specifically, we used transfer learning to extract patch level features followed by adding a max-pooling layer with only maximum feature values were kept along each feature dimension so that most irrelevant features that would potentially add noise to the model learning throughout the WSI were removed during this step. The features that were kept to train the top MLP were distributed widely throughout the whole slide image but not from a local region so that intra-tumor heterogeneity problem is reduced during training. We successfully demonstrated the effectiveness of this strategy by achieving high accuracy (=74.3%) in classifying genomic CIN status in test dataset.

Digital pathology images can be examined at different magnification levels. We experimented on both 2.5x and 10x magnified tiles for the predictions. We found that 2.5x achieved more accurate predictions marginally than 10x (2.5x AUC: 0.82, 10x AUC: 0.76; p-value=0.06) and multi-scaled model by combining 2.5x with 10x magnification features (2.5x AUC: 0.82, multi-scaled AUC: 0.81; p-value=0.59), although not statistically different tested by DeLong’s method (**Figure S3**). We reasoned that each tile on 2.5x level can capture more relevant features with a wider spatial view than high resolution tiles. While on 10x magnification level, tiles were more likely to be covered up by some ‘unknown’ irrelevant features. A similar observation was made in Coudray’s study [9] where analyzing 5x patches led to higher accuracy than 20x patches.

To validate the rationale of utilizing pathological slide images to infer genomic alternations, we performed differentially expressed gene analysis between CIN low and CIN high patients. The results revealed the impact of chromosomal instability in breast cancer including activating multiple pathways relating to cell cycle and mitosis (**Figure S8b**). As expected that mitotic alterations can be identified in slide images on high resolution views. Future studies may focus on training machine learning models to detect aberrant mitotic events directly from H&E slides. This will require a very large training set of such events that is currently not available.

## Material and Methods

### Dataset

Whole slide images along with clinical and genomic data were downloaded from The Cancer Genome Atlas (TCGA), project TCGA-BRCA using the TCGAbiolinks R package [22]. Only formalin-fixed paraffin-embedded (FFPE) diagnostic H&E stained histopathology slides from primary tumor sites were used for this study. After removing slides that lack magnification information, and/or slides with artefacts including tissue folding, air bubbles and out-of-focus regions, the cohort consisted of 1,010 patients with 1,070 whole slide images (WSI).

Fraction genome altered (FGA), which was defined as the ratio of *Sum of altered genome size/ Total genome size analyzed*, was calculated using whole exome sequencing based copy number variation (CNV) data on the same TCGA patients. Copy-number segments were downloaded from TCGA. Segments with log transformed mean copy number values larger than 0.2 or less than -0.2 [23, 24] were treated as altered segments, respectively. Based on examining the FGA distribution for all 1,010, patients, we labelled patients with FGA less than 0.3 as low CIN; those with FGA above 0.3 were labelled as high CIN.

### Image Preprocessing

First, a single overall region of interest (ROI) with dimension of 2,048×2,048 pixels on 2.5x magnification (4mpp) was determined by a sliding window approach from each whole slide image, which typically has dimension of about 8,000×4,500 pixels. A simple thresholding method was used to distinguish tissue from white space background on greyscale space. All pixels with value lower than 215 were treated as tissue, otherwise as background. The ROI window that contained the highest percentage of tissue was kept for further processing. The selected window was then split into 8×8 non-overlapping patches each with dimension of 256×256 pixels. Quality control on patch level was conducted using the following method. All patches with tissue percentage less than 80% or with significant blurriness, pen marks or folded tissues were deleted. Color normalization was then performed to reduce batch effects across different data sources (**Figure 1a**). We performed Reinhard normalization to transform the color characteristics to a desired standard defined by the mean and standard deviations of target image (TCGA-AN-A0FK) from the cohort using Python library of HistomicsTK [25]. Patients with more than one WSIs were treated as having one big WSI and can get more than 64 (8×8) patches depending on how many WSIs they have, but 64 qualified patches were randomly chosen to prevent over-representing those patients. Overall, after deleting all unqualified patches, we obtained median of 51 (IQR: 32, 61) qualified patches for each patient that to be used for transfer learning.

### Transfer Learning

#### (1)Feature Extraction

Instead of training CNN architectures from scratch, we used commonly used pre-trained models as feature extractors. We passed all patches (256×256 pixels) of each patient through Densenet-121, Densenet-169, Densenet-201 [26], Xception [27] and Resnet-50 [28] networks that were pre-trained on ImageNet [29] without top layers. Then we got a set of feature vectors for every patient with dimensions of m*n where m denotes number of patches of one patient and n implies number of features according to specific architecture that was used. For example, we got matrix of 32*1,024 for patch features with the patient who has 32 patches by using Densenet-121. To adapt patch level features to patient level labels and reduce the noise generated by intra-tumor heterogeneity, we applied a max-pooling layer on top of patch features and got patient level features with the same dimension for all patients (**Figure 1b**) [30].

#### (2)Train Multi-Layer Perceptron (MLP)

The whole cohort was randomly divided into training and validation set (n=858 patients, 85%) and hold-out testing set (n=152, 15%) without any overlap for both patients and images. Then the 858 patients were further split into training (730, 85%) and validation (128, 15%) set for tuning hyperparameters. We implemented multi-layer perceptron (MLP) which consists of several fully connected layers to take patient level features extracted by each of the CNN architectures mentioned above as input respectively (**Figure 1c**). Therefore, output by each model will be a prediction probability of high CIN class patient. Our model used binary cross-entropy loss function. We initialized the MLP network weights with He initialization [31]. Adam optimizer was used for the training network weights with learning rate of 0.00001. Training was stopped early [32] if validation loss was not improving within 200 epochs. Epoch numbers were selected as per lowest validation loss. Then model performance metrics including ROC curve and AUC, balanced accuracy, sensitivity and specificity were calculated in the hold-out test set for the final evaluation (**Figure 1d**).

### Visualization of predicted patches

The trained MLP from last step can be fed with both combined patches (patient level features) and individual patches (patch level features) for hierarchical predictions. We predicted both patient level and patch level high CIN probabilities and visualized the predictions to demonstrate the existence of intra-tumor heterogeneity (**Figure 1e**).

### Pathological, Genomic and Transcriptomic scoring of CIN

We define predicted CIN-high fraction score as the percentage of predicted high CIN patches based on each pathology slide image. CIN-max, CIN-95^th^ and CIN-75^th^ are defined as maximum, 95^th^ percentile and 75^th^ percentile of patch predictions within each slide, respectively. Genomic CIN score was calculated by fraction genome altered as mentioned earlier [24]. CIN23 gene signature [4] score was computed using single sample Gene Set Enrichment Analysis (ssGSEA) to indicate transcriptional CIN score from RNA expression data of the same cohort.

### Transcriptome analysis

We examined differentially expressed genes of primary tumor between high CIN and low CIN patients using Limma R package [33]. Differential expressed genes and samples shown in heatmap were clustered using Euclidean distance. Gene set enrichment analysis was conducted using fgsea [34] R package and Reactome pathway database (https://reactome.org).

### Mitosis events inspection

Among all the patients who have agreed predicted CIN and genomic CIN status, top five extreme high CIN and low CIN patients were selected according to genomic CIN score. Several tumor patches of one patient with dimension of 1,024×1,024 on 40x magnification were randomly inspected and atypical mitosis events number in each patch were recorded.

### Statistical analysis and Software

Training of our DNN method was performed on local computer powered by one NVIDIA GeForce GT 640M GPU with 512 MB memory and one 2.7-GHz Quad-Core Intel Core i5 CPU. All statistical and bioinformatics analyses were performed in R, version 3.6.2. Image preprocessing and neural network training were conducted in Python, version 3.7.4. ROC curves were compared using DeLong’s method by R package of pROC. Chi-square test was conducted to test the independency between cancer subtypes with genomic CIN status. Spearman’s rank-order correlation test was performed for the correlation analysis without distribution assumption. Log rank test was used for comparing Kaplan-Meier survival curves between different genome and predicted CIN groups. Time to any new tumor events and mortality was used as composite survival events and the data was censored at 5 years. Maximally selected rank statistics [35, 36] was used to determine the optimal prognostic cutoff points for CIN biomarkers including predicted CIN, predicted CIN-High fraction, CIN-max, CIN-95^th^ and CIN-75^th^. Generalized estimating equation (GEE) of Poisson regression model was used to compare atypical mitosis event number between CIN high and CIN low groups. All statistical tests were two sided with p<0.05 indicated significant. OpenSlide python was used for reading and tiling whole-slide images. TensorFlow2 were used for building and training neural networks. The source code and the guideline are publicly available at https://github.com/eipm/CIN.

## Data Availability

Whole slide images along with clinical and genomic data can be downloaded from The Cancer Genome Atlas (TCGA), project TCGA-BRCA. The source code and the guideline will be publicly available at https://github.com/eipm/CIN.

## References

[1] C. Lengauer, K. W. Kinzler and B. Vogelstein, “Genetic instabilities in human cancers,” Nature, vol. 396, pp. 643–649, 17 12 1998.

[2] L. Sansregret, B. Vanhaesebroeck and C. Swanton, “Determinants and clinical implications of chromosomal instability in cancer,” Nature Reviews, vol. 15, pp. 139–148, 2018.

[3] S. F. Bakhoum and L. C. Cantley, “The multifaceted role of chromosomal instability in cancer and its microenvironment,” Cell, vol. 147, no. 6, pp. 1347–1360, 06 09 2018.

[4] S. F. Bakhom, “Chromosomal instability drives metastasis through a cytosolic DNA response,” Nature, vol. 553, pp. 467–472, 2018.

[5] M. Smid and M. Hoes, “Patterns and incidence of chromosomal instability and their prognostic relevance in breast cancer subtypes,” Breast Cancer Research and Treatment, vol. 128, pp. 23–30, 2011.

[6] S. L. Carter and A. C. Eklund, “A signature of chromosomal instability inferred from gene expression profiles predicts clinical outcome in multiple human cancers,” Nature Genetics, no. 38, pp. 1043–1048, 2006.

[7] L. M. Zasadil, K. A. Andersen and D. Yeum, “Cytotoxicity of Paclitaxel in Breast Cancer Is Due to Chromosome Missegregation on Multipolar Spindles,” Science Translational Medicine, vol. 6, no. 229, 26 03 2014.

[8] D. D. Pierssens, M. C. Borgemeester and S. J. van der Heijden, “Chromosome instability in tumor resection margins of primary OSCC is a predictor of local recurrence,” Oral Oncology, vol. 66, pp. 14–21, March 2017.

[9] N. Coudray, A. L. Moreira and T. Sakellaropoulos, “Classification and Mutation Prediction from Non-Small Cell Lung Cancer Histopathology Images using Deep Learning,” Nature Medicine, vol. 24, pp. 1559–1567, 2018.

[10] A. J. Schaumberg, M. A. Rubin and T. J. Fuchs, “H&E-stained Whole Slide Image Deep Learning Predicts SPOP Mutation State in Prostate Cancer,” bioRxiv 064279; doi: https://doi.org/10.1101/064279, 2016.

[11] H. Xu, S. Park, S. H. Lee and T. H. Hwang, “Using transfer learning on whole slide images to predict tumor mutational burden in bladder cancer patients,” bioRxiv 554527; doi: https://doi.org/10.1101/554527, 2019.

[12] J. N. Kather, A. T. Pearson and N. Halama, “Deep learning can predict microsatellite instability directly from histology in gastrointestinal cancer,” Nature Medicine, vol. 25, pp. 1054–1056, July 2019.

[13] J. N. Kather, L. R. Heij and T. Luedde, “Pan-cancer image-based detection of clinically actionable genetic alterations,” Nature Cancer, vol. 1, pp. 789–799, 2020.

[14] Y. Fu, A. W. Jung and R. V. Torne, “Pan-cancer computational histopathology reveals mutations, tumor composition and prognosis.,” Nature Cancer, vol. 1, pp. 800–810, 2020.

[15] J. D. Schonhoft, J. L. Zhao and A. Jendrisak, “Morphology-predicted large scale transition number in circulating tumor cells identifies a chromosomal instability biomarker associated with poor outcome in castration-resistant prostate cancer,” Cancer Research, 2020. DOI: 10.1158/0008-5472.CAN-20-1216

[16] K. A. Kwei and Y. Kung, “Genomic instability in breast cancer: pathogenesis and clinical implications,” Molecular Oncology, vol. 4, pp. 255–266, 2010.

[17] P. Khosravi, E. Kazemi, M. Imielinski, O. Elemento and I. Hajirasouliha, “Deep convolutional neural networks enable discrimination of heterogeneous digital pathology images,” EBioMedicine, no. 27, pp. 317–328, 2018.

[18] B. Orsetti, J. Selves and C. Bascoul-Mollevi, “Impact of chromosomal instability on colorectal cancer progression and outcome,” BMC Cancer, vol. 1, no. 14, pp. 1–13, 2014.

[19] N. J. Birkbak and A. C. Eklund, “Paradoxical Relationship between Chromosomal Instability and Survival Outcome in Cancer,” Cancer Research, vol. 71, no. 10, pp. 3447–3452, 2011.

[20] L. M. Zasadil and E. M. Britigan, “High rates of chromosome missegregation suppress tumor progression but do not inhibit tumor initiation,” Molecular Biology of the Cell, vol. 27, no. 13, pp. 1981–2144, 2016.

[21] C. L. Srinidhi, O. Ciga and A. L. Martel, “Deep neural network models for computational histopathology: A survey,” ArXiv, 2019.

[22] A. Colaprico, “TCGAbiolinks: An R/Bioconductor package for integrative analysis of TCGA data,” Nucleic Acids Research, 2015.

[23] L. A. Salas and K. C. Johnson, “Integrative epigenetic and genetic pan-cancer somatic alteration portraits,” vol. 12, no. 7, pp. 561–574, 2017.

[24] N. Z. Ali Hassan and N. M. Mokhtar, “Integrated analysis of copy number variation and genome-wide expression profiling in colorectal cancer tissues,” Plos One, vol. 9, no. 4, 2014.

[25] D. A. Gutman and M. Khalilia, “The digital slide archive: a software platform for management, integration and analysis of histology for cancer research,” Cancer Research, vol. 77, pp. e75–e78, 2017.

[26] G. Huang and Z. Liu, “Densely Connected Convolutional Networks,” IEEE Conference on Pattern Recognition and Computer Vision (CVPR), 2017.

[27] F. Chollet, “Xception: Deep Learning with Depthwise Separable Convolutions,” IEEE Conference on Computer Vision and Pattern Recognition (CVPR), 2017.

[28] K. He and X. Zhang, “Deep Residual Learning for Image Recognition,” IEEE Conference on Computer Vision and Pattern Recognition (CVPR), 2016.

[29] J. Deng, “ImageNet: A large-scale hierarchical image database.,” IEEE Conference on Computer Vision and Pattern Recognition, 2009.

[30] P. Courtiol and E. W. Tramel, “Classification and disease Localization in histopathology using only global labels: A weakly-supervised approach,” arXiv, 2018.

[31] K. He, X. Zhang, S. Ren and J. Sun, “Delving Deep into Rectifiers: Surpassing Human-Level Performance on ImageNet Classification,” ICCV, pp. 1026-1034, 2015.

[32] L. Prechelt, “Early Stopping -But When?.,” in Neural Networks: Tricks of the Trade, Springer, 2012, pp. 53–67.

[33] M. E. Ritchie and B. Phipson, “Limma powers differential expression analyses for RNA-sequencing and microaary studies,” Nucleic Acids Research, vol. 43, no. 7, p. e47, 2015.

[34] G. Korotkevich, V. Sukhov and A. Sergushichev, “Fast gene set enrichment analysis,” bioRxiv, 2019.

[35] B. Lausen, T. Hothorn, F. Bretz and M. Schumacher, “Assessment of Optimal Selected Prognostic Factors,” Biomedical Journal, vol. 46, no. 3, pp. 364–374, 2004.

[36] T. Hothorn and B. Lausen, “On the exact distribution of maximally selected rank statistics,” Computational Statistics & Data Analysis, vol. 43, no. 2, pp. 121–137, 2003.

[37] E. Montgomery, R. E. Wilentz and P. Argani, “Analysis of Anaphase Figures in Routine Histologic Sections Distinguishes Chromosomally Unstable from Chromosomally Stable Malignancies,” Cancer Biology & Therapy, vol. 2, no. 3, pp. 248–252, 2003.

[38] Y. Matsuda and J. Aida, “Morphological Markers of Chromosomal Instability,” in Chromosomal Abnormalities: A Hallmark Manifestation of Genomic Instability, IntechOpen, 2017.

[39] R. Cailleau, M. Olive and Q. V. Cruciger, “Long-term human breast carcinoma cell lines of metastatic origin: Preliminary characterization,” In Vitro, vol. 14, pp. 911–915, 1978.

